# Assessment of Hygienic Practices in Street Food Vendors of Mymensingh City of Bangladesh: A Cross-Sectional Study

**DOI:** 10.64898/2026.03.27.26349369

**Authors:** FNU Nahiduzzaman, Tasnim Zarin, Nishat Ara Jhinuk, Hasebul Hasan, Mst. Minara Khatun, Md. Ariful Islam

## Abstract

This cross-sectional study assessed hygienic practices, microbial contamination, and associated factors among street food vendors in Mymensingh City, Bangladesh, from August 2024 to February 2025. Using purposive sampling, 300 vendors were evaluated through structured questionnaires, observational assessments, and laboratory analysis of food samples (n=300) for bacterial load (log-transformed Total Viable Count, log_TVC). Results revealed that 87.33% (95% CI: 83.6-91.1) of vendors practiced poor hygiene: 90.7% (95% CI: 87.4-94.0) did not cover food, 7% (95% CI: 4.1-9.9) used disinfectants, and 81.00% (95% CI: 76.6-85.4) reused ingredients beyond one day. Knowledge gaps were stark, with 89.7% (95% CI: 86.3-93.1%) demonstrating insufficient basic hygiene knowledge and 90.7% (95% CI: 87.4-94.0%) lacking understanding of hygienic food processing. Education significantly influenced practices; secondary-educated vendors exhibited markedly better hygiene awareness (65.0% vs. 2.89% in uneducated groups). Bacterial loads varied across food types, with Velpuri showing the highest mean log_TVC (8.00, 95% CI: 7.65-8.35) and Fast food the lowest (7.69, 95% CI: 7.34-8.03). Significant correlations emerged between hygiene knowledge and practices: vendors aware of foodborne diseases were more likely to use gloves (Spearman’s r=0.199, p=0.001) and cover food (r=0.118, p=0.041). Challenges included financial constraints (62.25% [95% CI: 56.1-68.4] among uneducated vendors), knowledge gaps, and time limitations. The study underscores systemic issues in street food safety, driven by low education and economic barriers. Interventions targeting vendor education, affordable hygiene solutions, and stricter enforcement of ingredient freshness protocols are urgently needed to mitigate foodborne disease risks in urban Bangladesh.

## 1. Introduction

Street food vending is a key component of urban life, providing accessible and affordable meal options to large part of the population, particularly in developing countries like Bangladesh. The informal food sector is essential for food security among low- and middle-income populations and provides a significant source of livelihood for many vendors. (Uddin, 2021). Street food’s low cost, convenience, and variety make it a key meal source for students, workers, and low-income families (Ferrari et al., 2021). Moreover, street food vending offers self-employment opportunities for individuals, especially those with limited formal education or access to the formal job market (Giraldo et al., 2020). Street food vendors often operate in busy areas like markets, transport hubs, and schools, offering ready-to-eat items such as fuchka, chotpoti, velpuri, fruits, fast foods, and drinks (Al Mamun et al., 2013; Husain et al., 2015; Uddin, 2021). Although street foods support the local economy and provide affordable meals, their informal nature often makes maintaining proper food safety standards difficult (Uddin, 2021). Since vendors typically operate without formal training in food hygiene and lack access to proper infrastructure, their food handling practices may not meet standard safety regulations (Islam et al., 2024). Consequently, street-vended foods are frequently linked to hygiene concerns, raising considerable public health risks (Nahiduzzaman et al., 2025). Foodborne illnesses are a major global health concern, with low- and middle-income countries being disproportionately affected. The World Health Organization estimates that about 600 million people globally develop foodborne illnesses each year, causing roughly 420,000 deaths. (Kang and White, 2016). Contaminated street foods are a major source of foodborne illness including diarrhea, fever, vomiting and other gastrointestinal symptoms that are subjected to food poisoning (Nahiduzzaman et al., 2025). The presence of bacterial pathogens such as *Escherichia coli*, *Salmonella spp.*, and *Staphylococcus aureus* has frequently been reported in contaminated street foods in Bangladesh (Banik et al., 2018; Samad et al., 2023; Nahiduzzaman et al., 2025). In Bangladesh, outbreaks of foodborne illnesses have been widely reported, with contaminated street-vended foods being a common factor (Afzalur Rahman et al., 2018; Salamandane et al., 2023). Such incidents often arise from poor food handling, inadequate sanitation practices, and improper food storage conditions. The informal nature of street vending makes regulatory oversight particularly challenging, further increasing the risk of food contamination (Rakha et al., 2022). As a result, contaminated food may enter the market unnoticed, posing significant health risks to consumers. The hygiene practices of street food vendors are influenced by various factors, including their knowledge, attitudes, and environmental conditions (Banna et al., 2022). Vendors with limited awareness of food safety principles may unintentionally adopt unsafe practices, such as infrequent handwashing, using unclean utensils, vendors operating outdoor places or handling raw and cooked foods with the same tools (Farhana et al., 2020; Wallace et al., 2022). Inadequate waste disposal systems near vending sites also contribute to unhygienic conditions, providing an environment conducive to bacterial growth (Nkosi and Tabit, 2021). Financial limitations may prevent vendors from purchasing essential protective gear such as gloves, aprons, or food covers (Kundu et al., 2021; Islam et al., 2024). As a result, most street vendors in Bangladesh often resort to unsafe practices, like reusing contaminated utensils, preparing food on unclean surfaces, or storing food in unsanitary conditions (Hassan et al., 2017). These practices increase the likelihood of food contamination, placing consumers at heightened risk of illness. While some Bangladeshi studies have assessed street food microbiology, research on vendors’ hygiene, food safety knowledge, and contamination risks in Mymensingh City is scarce. With growing dependence on street foods, such evaluation is crucial to identify risks and guide interventions.

This study assesses street food vendors in Mymensingh City, examining their personal hygiene, food handling, and sanitation, alongside their food safety knowledge, attitudes, and challenges. It also explores links between hygiene practices and bacterial contamination, aiming to inform targeted interventions for improving food safety in Bangladesh’s informal food sector.

## 2. Methodology

### 2.1 Study Design and Sampling

This study was conducted in Mymensingh City (Around an average radius of 8 kilometers centered at 24.749483°N, 90.399321°E), Bangladesh, from August 2024 to February 2025. A cross-sectional study design was employed to evaluate the hygienic practices of street food vendors. A total of 300 vendors were selected using a purposive sampling technique. The selection criteria included vendors who had been operating for at least six months, sold ready-to-eat food items, and were willing to participate in the study.

### 2.2 Food Sample Collection and Analysis

Food samples (50 g) were collected from each vendor and transported to the laboratory in ice boxes to maintain sample integrity. Each sample was homogenized using a grinder by mixing with 50 ml of 0.1% peptone water (HiMedia, India). Following homogenization, a 10-fold serial dilution was performed up to a 10 ¹ dilution. From each diluted sample, 100 µl was plated using the Spread Plate Technique on Plate Count Agar (HiMedia, India). The plates were incubated at 37°C for 18-24 hours, after which the number of bacterial colonies was counted. The Colony Forming Units (CFU) were calculated using the standard formula described by (Hasan et al., 2023) and expressed as log_TVC values for consistency in bacterial load comparison.

### 2.3 Data Collection

Data was collected through face-to-face interviews using a structured questionnaire on demographics, business details, and food hygiene knowledge, combined with direct observations of food handling, personal hygiene, storage, water use, and cleaning practices. The study examined vendors’ hygiene behaviors, awareness of foodborne diseases, and challenges such as financial, time, and knowledge constraints.

### 2.4 Data Analysis and Statistical Tests

Descriptive statistics summarized vendor demographics, hygiene practices, and food safety knowledge, while Chi-square tests and Cramér’s V evaluated associations between education, experience, and hygiene behaviors (McHugh, 2013). Cramér’s V was calculated using the formula:

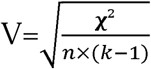

where χ**²** is the Chi-square statistic, ***n*** is the sample size, and ***k*** is the smaller number of variable categories. Values range from 0 (no association) to 1 (perfect association). Interpretation thresholds were: 0–0.1 (negligible), 0.1–0.3 (weak), 0.3–0.5 (moderate), and >0.5 (strong). Associations between hygiene practices and bacterial load were evaluated using Spearman’s rank and Pearson’s correlation tests, while the Kruskal-Wallis test compared median log_TVC values across food types (McKight and Najab, 2010; Hauke and Kossowski, 2011). A p-value < 0.05 was considered statistically significant, with correlation strengths interpreted as follows: 0.10–0.29 (weak), 0.30–0.49 (moderate), and ≥0.50 (strong). All analyses were conducted using SPSS (v27) and PyCharm (Python 3.14).

## 3. Results

### 3.1 Demographic Information of the Street Food Vendors

The demographic distribution of street food sellers shows that the majority fall within the 19–35 years age group (40.0%; 95% CI: 34.5–45.5%), followed by the 36–50 years cohort (31.3%; 95% CI: 26.1–36.6%) and the 50–70 years group (28.3%; 95% CI: 23.2–33.4%). Only 0.3% (95% CI: 0.05–1.67%) are under 18. Males dominate the workforce (96.0%; 95% CI: 93.8– 98.2%), with females representing a small minority (4.0%; 95% CI: 1.8–6.2%). Educational attainment is strikingly limited: 80.7% (95% CI: 76.2–85.2%) are uneducated, 12.7% (95% CI: 8.9–16.4%) have primary education, and 6.7% (95% CI: 3.9–9.5%) possess secondary education. The most prevalent food items sold are Fuchka and Chotpoti (29.3%; 95% CI: 24.2–34.4%), followed by fruits (15.7%; 95% CI: 11.6–19.8%), Jhalmuri (15.0%; 95% CI: 11.0–19.0%), and Fuchka with Velpuri (12.0%; 95% CI: 8.3–15.7%). Less common offerings include Velpuri (9.7%; 95% CI: 6.3–13.1%), Fuchka alone (7.3%; 95% CI: 4.4–10.2%), fast food (6.7%; 95% CI: 3.9–9.5%), and Chotpoti (4.3%; 95% CI: 2.0–6.6%).

### 3.2 Knowledge of Foodborne Diseases and Hygiene

The study revealed critical deficiencies in hygiene-related knowledge among street food sellers. Insufficient basic hygiene knowledge (BKH) was observed in 89.7% (95% CI: 86.3–93.1%) of sellers, while 82.0% (95% CI: 77.7–86.3%) lacked awareness of foodborne diseases (KFD). Alarmingly, 90.7% (95% CI: 87.4–94.0%) demonstrated an inadequate understanding of hygienic food processing (KHFP), with no respondents achieving “sufficient” proficiency in any category. Striking educational disparities emerged: secondary-educated sellers displayed significantly higher competency in moderately sufficient BKH (65.0% [95% CI: 55.1–74.9%]), KFD awareness (90.0% [95% CI: 83.2–96.8%]), and KHFP understanding (95.0% [95% CI: 89.8–100%]) compared to uneducated peers (BKH: 2.89% [95% CI: 0.85–4.93%], KFD: 5.79% [95% CI: 2.8–8.8%], KHFP: 0.83% [95% CI: 0.0–2.1%]) as shown in (Figure 1). These results highlight education as a critical determinant of food safety practices (χ²=132.7, p<0.001).

**Figure 1:**
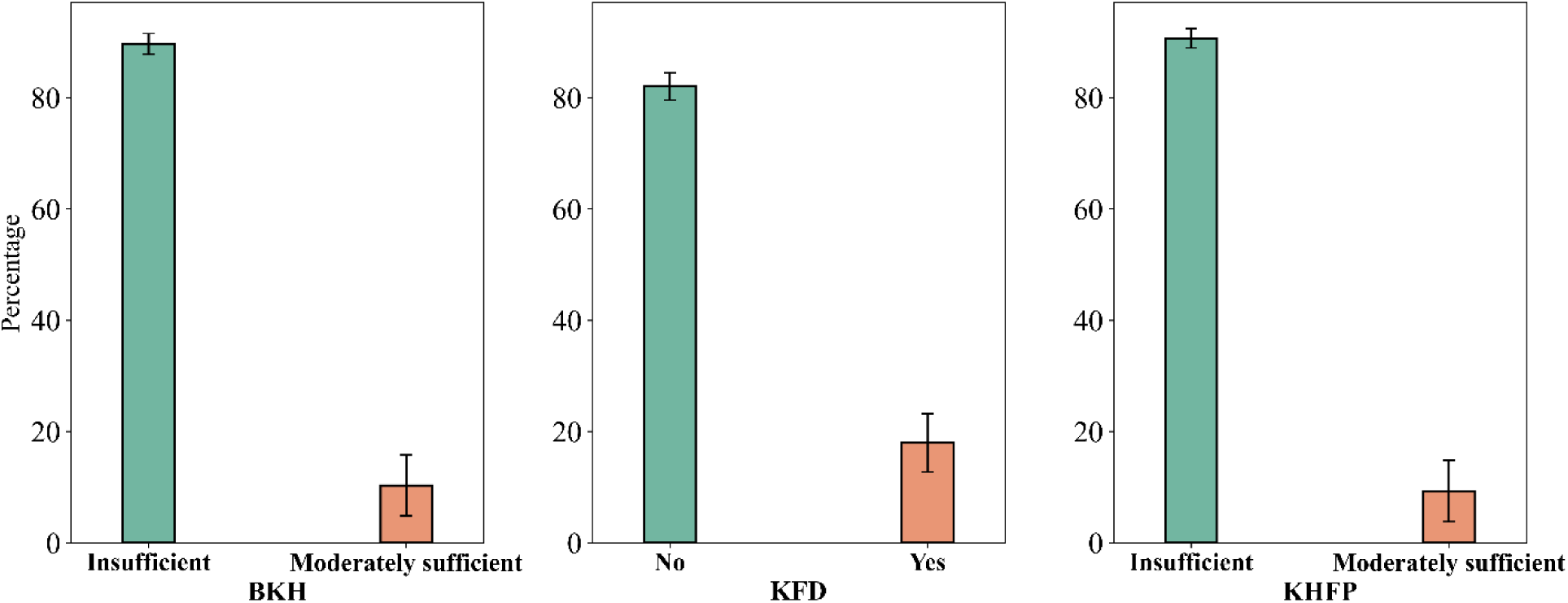
Status of knowledge about basic hygiene, hygienic food processing and foodborne diseases

### 3.3 Hygienic Practices in Street Food Vending

The study revealed concerning hygiene practices among street food vendors in Mymensingh city. A significant majority, 87.33% (95% CI: 83.6–91.1), of vendors did not adhere to hygienic food-selling practices, with only 12.7% (95% CI: 8.9–16.4) following appropriate measures. Most vendors, 95.3% (95% CI: 92.9–97.7), prepared food at their selling sites rather than at home, 4.7% (95% CI: 2.3–7.1), posing a higher risk of contamination. Alarmingly, 90.7% (95% CI: 87.4–94.0) of vendors did not cover their food, while only 9.3% (95% CI: 6.0–12.6) ensured food protection. Water hygiene was also inadequate, with 81.3% (95% CI: 76.9–85.7) using unhygienic water, 10% (95% CI: 6.6–13.4) using hygienic water, and 8.7% (95% CI: 5.5–11.9) using moderately hygienic sources. Disinfectant use was minimal, with only 7% (95% CI: 4.1–9.9) employing it during food preparation. Similarly, glove usage was rare, with 90.3% (95% CI: 87.0–93.7) of vendors neglecting this practice. Hand hygiene practices were also inadequate, with 81.3% (95% CI: 76.9–85.7) washing their hands insufficiently, 15.3% (95% CI: 11.3–19.4) moderately sufficiently, and only 3.3% (95% CI: 1.3–5.4) sufficiently. The cleaning frequency of utensils showed similar trends, with 81% (95% CI: 76.6–85.4) of vendors cleaning utensils insufficiently, 10.7% (95% CI: 7.2–14.2) moderately sufficiently, and 8.3% (95% CI: 5.2–11.5) sufficiently. Regarding food storage, 81% (95% CI: 76.6–85.4) reused the same food ingredients for more than one day, while only 19% (95% CI: 14.6–23.4) utilized fresh ingredients daily. These findings in (Figure 2) underscore the urgent need for improved hygiene awareness and regulatory measures to ensure safer street food practices.

**Figure 2:**
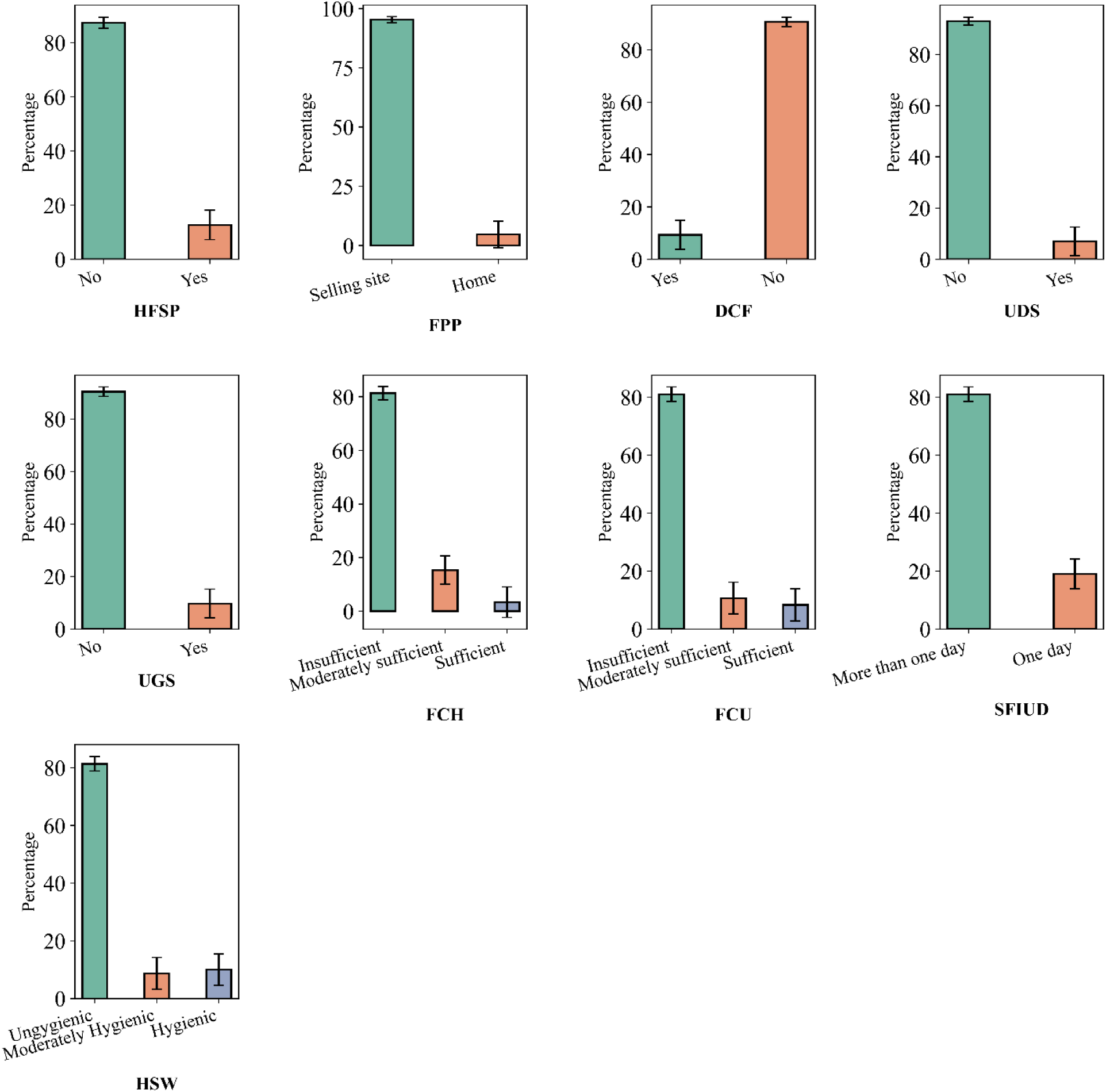
Status of hygienic practices during street food preparation and selling

### 3.4 Bacterial load analysis

The analysis of the log-transformed Total Viable Count (TVC) for various food categories reveals distinct differences in bacterial loads across food types. Chotpoti has a mean log_TVC of 7.95 (95% CI: 7.52, 8.39), while Fast food shows a mean log_TVC of 7.69 (95% CI: 7.34, 8.03).

Fruits has a mean log_TVC of 7.75 (95% CI: 7.51, 7.98), and Fuchka displays a mean log_TVC of 7.92 (95% CI: 7.51, 8.33). The combination of Fuchka & Chotpoti has a mean log_TVC of 7.85 (95% CI: 7.66, 8.05), while Fuchka & Velpuri shows a mean log_TVC of 7.96 (95% CI: 7.65, 8.27). Jhalmuri has a mean log_TVC of 7.75 (95% CI: 7.48, 8.02). The highest mean log_TVC, 8.00, is observed in Velpuri (95% CI: 7.65, 8.35). These results, as shown in (Figure 3) suggest that bacterial loads vary among different food types, with Velpuri showing the highest mean, while Fast food exhibits the lowest mean log_TVC.

**Figure 3:**
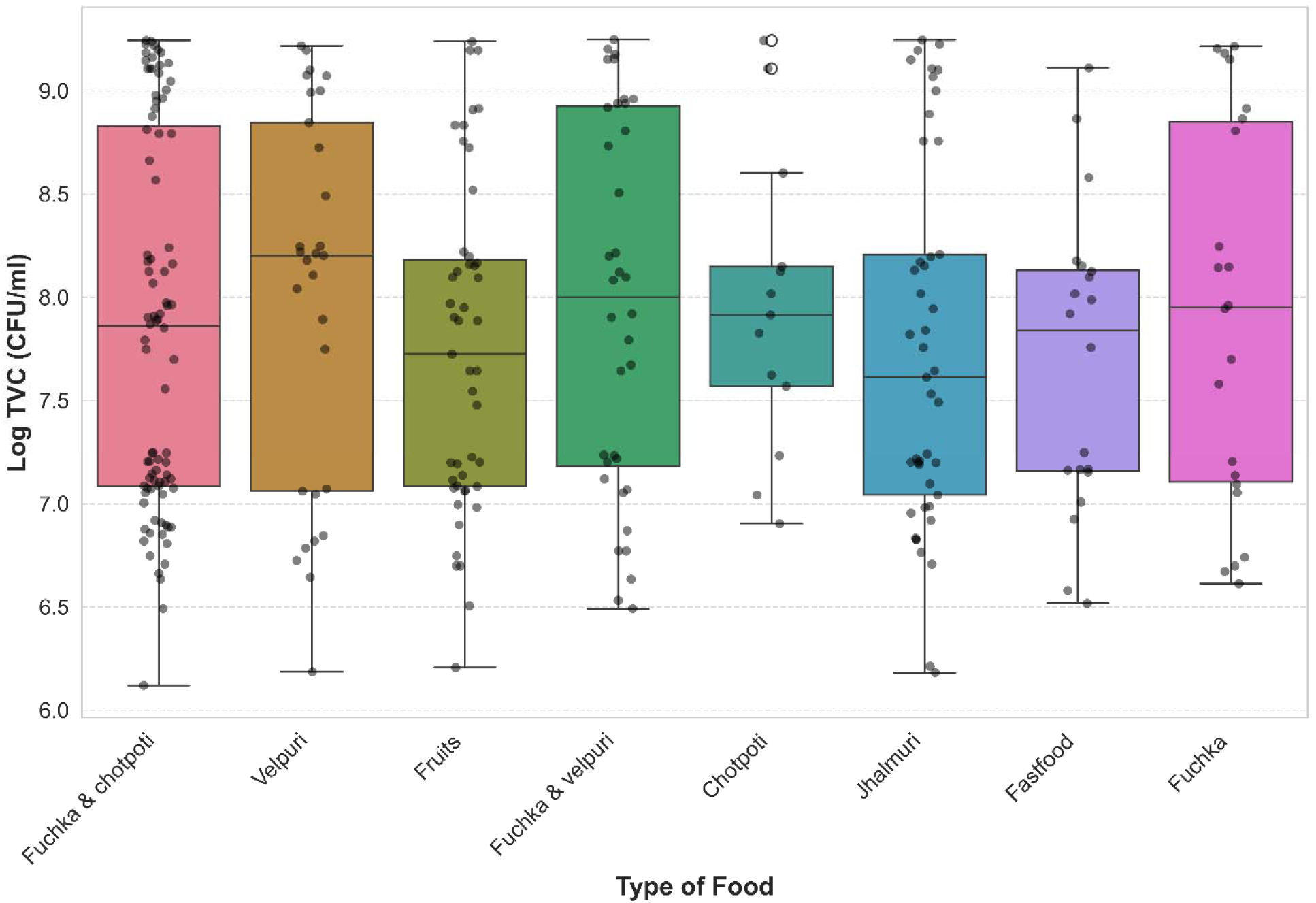
Bacterial load for different food samples collected from the street vendors

### 3.5 Factors Affecting Hygienic Practices

The analysis of data from 300 street food sellers revealed noteworthy associations between knowledge and hygienic practices. Spearman’s rank and Pearson’s correlation coefficients indicated several significant correlations, particularly between “Knowledge about foodborne diseases” (KFD) and practices such as “Using gloves during food selling” (DUG), which showed moderate positive correlations (Spearman’s r = 0.199, p = 0.001; Pearson’s r = 0.199, p = 0.001). This suggests that vendors with a better understanding of foodborne diseases are more likely to adopt proper hygiene practices. Similarly, “Basic knowledge of hygiene” (BKH) showed a positive but weak correlation with “Hygienic food selling practices” (HFSP), though not statistically significant (Spearman’s r = 0.155, p = 0.007). “Knowledge about foodborne diseases” (KFD) was significantly correlated with “Proper food covering” (DCF), with a moderate positive relationship observed (Spearman’s r = 0.118, p = 0.041), indicating that vendors knowledgeable about contamination risks are more likely to practice food covering. Furthermore, “Knowledge about hygienic food processing” (KHFP) showed a weak but significant correlation with “Same food ingredients use (in days)” (SFIUD), with a Spearman’s r of 0.171 (p = 0.003), suggesting that education on disinfection could enhance the use of disinfectants in food-selling environments.

The chi-square analysis further supported these findings, revealing significant associations between knowledge variables and practices, particularly in the use of disinfectants (χ² = 9.055, p = 0.003) and proper food covering (χ² = 5.530, p = 0.019). Cramer’s V analysis indicated moderate to strong associations between these variables, with KFD correlating with hygienic practices at a meaningful level (Cramer’s V = 0.174 for HFSP and KFD). Additionally, KFD showed a moderate association with “Proper food covering” (DCF), with a Cramer’s V value of 0.160, suggesting that increased awareness of foodborne diseases leads to better food safety practices, such as covering food appropriately. These results underscore the pivotal role of hygiene knowledge in shaping food safety behaviors, suggesting that educational interventions focused on foodborne diseases and proper hygiene could significantly enhance hygienic practices among street food vendors (Figure 4) and supplementary (Table 1).

**Figure 4.**
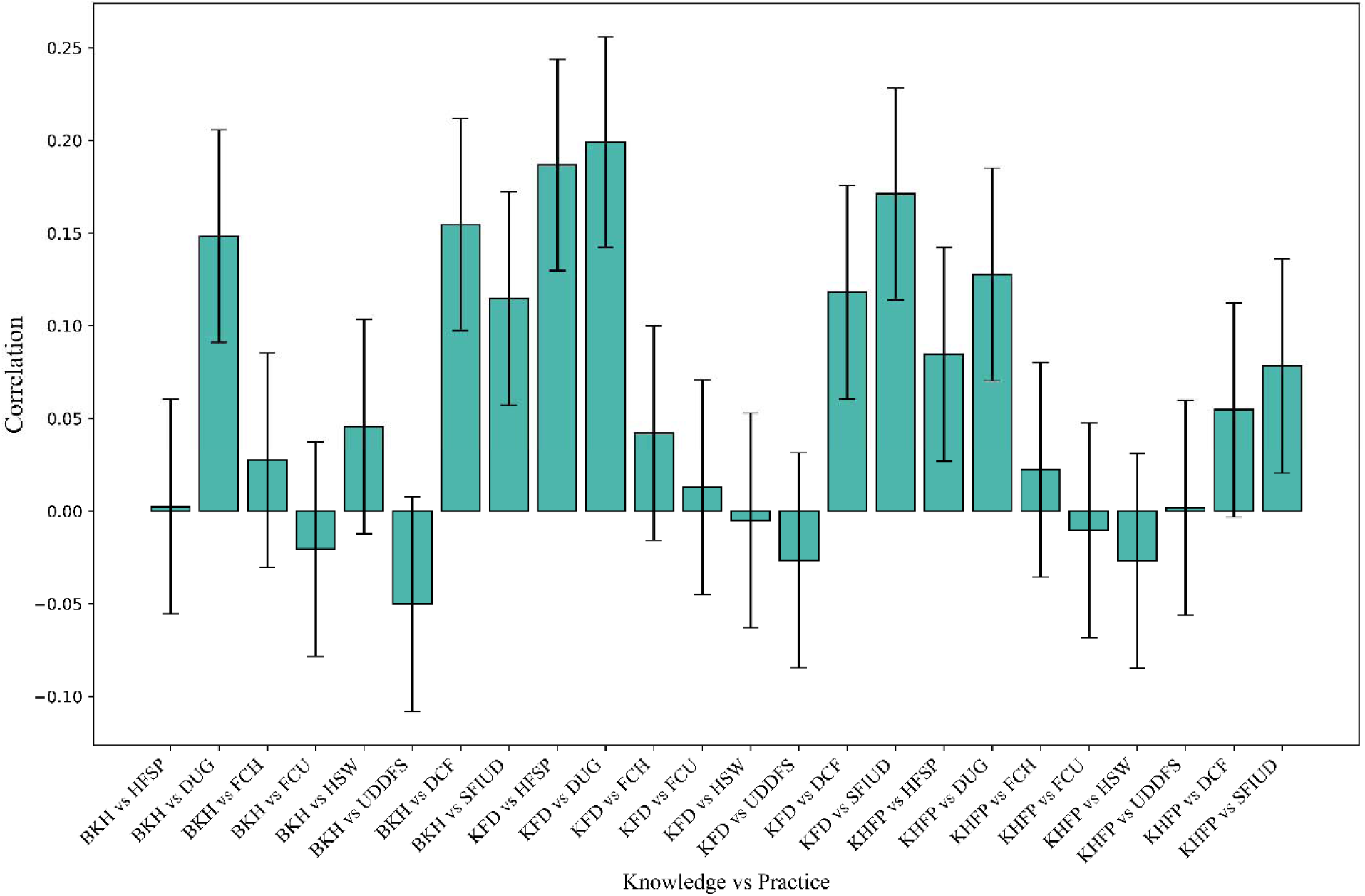
Spearman’s rank correlation coefficients (± standard error) illustrating the associations between different domains of food safety knowledge and hygienic practices in street food vendors.

**Table 1:** Abbreviations used during statistical analysis.

| Abbreviation | Full Term |
| --- | --- |
| BKH | Basic knowledge of hygiene |
| KFD | Knowledge about foodborne diseases |
| KHFP | Knowledge about hygienic food processing |
| HFSP | Hygienic food selling practices |
| DCF | Does he cover the food |
| UDDFS | Uses of Disinfectant during food selling |
| UGS | Does he use gloves during food selling |
| FCH | Frequency of cleaning hands |
| FCU | Frequency of cleaning utensils |
| SFIUD | Same food ingredients use (in days) |
| HSW | Hygienic status of water |
| DUG | Does he use gloves during food selling |

### 3.6 Correlation of Hygienic Practices and Bacterial Load

The comprehensive analysis revealed critical associations between hygiene practices and microbial contamination (log-TVC) across street food categories. Fuchka & chotpoti vendors exhibited a significant negative correlation between Same food ingredients use (in days) (SFIUD) and microbial load (r = -0.21, p = 0.046, n = 88), indicating a 21% reduction in contamination with daily ingredient replacement. Velpuri sellers demonstrated improved safety outcomes tied to the Frequency of cleaning utensils (FCU) (r = 0.39, p = 0.037, n = 29), while Fuchka vendors showed a robust protective effect of SFIUD (r = -0.49, p = 0.021, n = 22), halving contamination risks with fresh ingredients. Population-level Kruskal-Wallis tests confirmed systemic microbial load differences linked to SFIUD (H = 14.10, p < 0.001, N = 300), with median log-TVC values of 1.8 CFU/ml higher among vendors reusing ingredients beyond one day. Marginally significant trends included the Hygienic Status of Water (HSW) for Fast food (r = -0.43, p = 0.062, n = 20), suggesting better water quality may reduce contamination, and Uses of Disinfectant (UDDFS) for Fuchka (r = 0.40, p = 0.066, n = 22), highlighting potential benefits of sanitization. Notably, HSW showed category-specific trends: Velpuri (r = 0.35, p = 0.059, n = 29) and Chotpoti (r = 0.17, p = 0.577, n = 13), though not statistically significant. Contrary to assumptions, Food Preparation Place (FPP) had no measurable impact (p = 0.955, N = 300), challenging the notion that home-based preparation inherently improves safety. Similarly, Frequency of Cleaning Hands (FCH) and Glove Usage (DUG) showed no significant correlations across categories (p > 0.05). These findings, as shown in (Figure 5), (Table 2) and supplementary (Table 2 & 3), underscore SFIUD as the most critical factor for contamination control, with a population-wide effect size (η² = 0.047) explaining 4.7% of microbial load variance. Targeted strategies are needed since FCU affects Velpuri most. Regulations should focus on daily ingredient turnover and standardized utensil cleaning, while water quality and disinfectant use in high-risk items like Fast food and Fuchka require further study.

**Figure 5:**
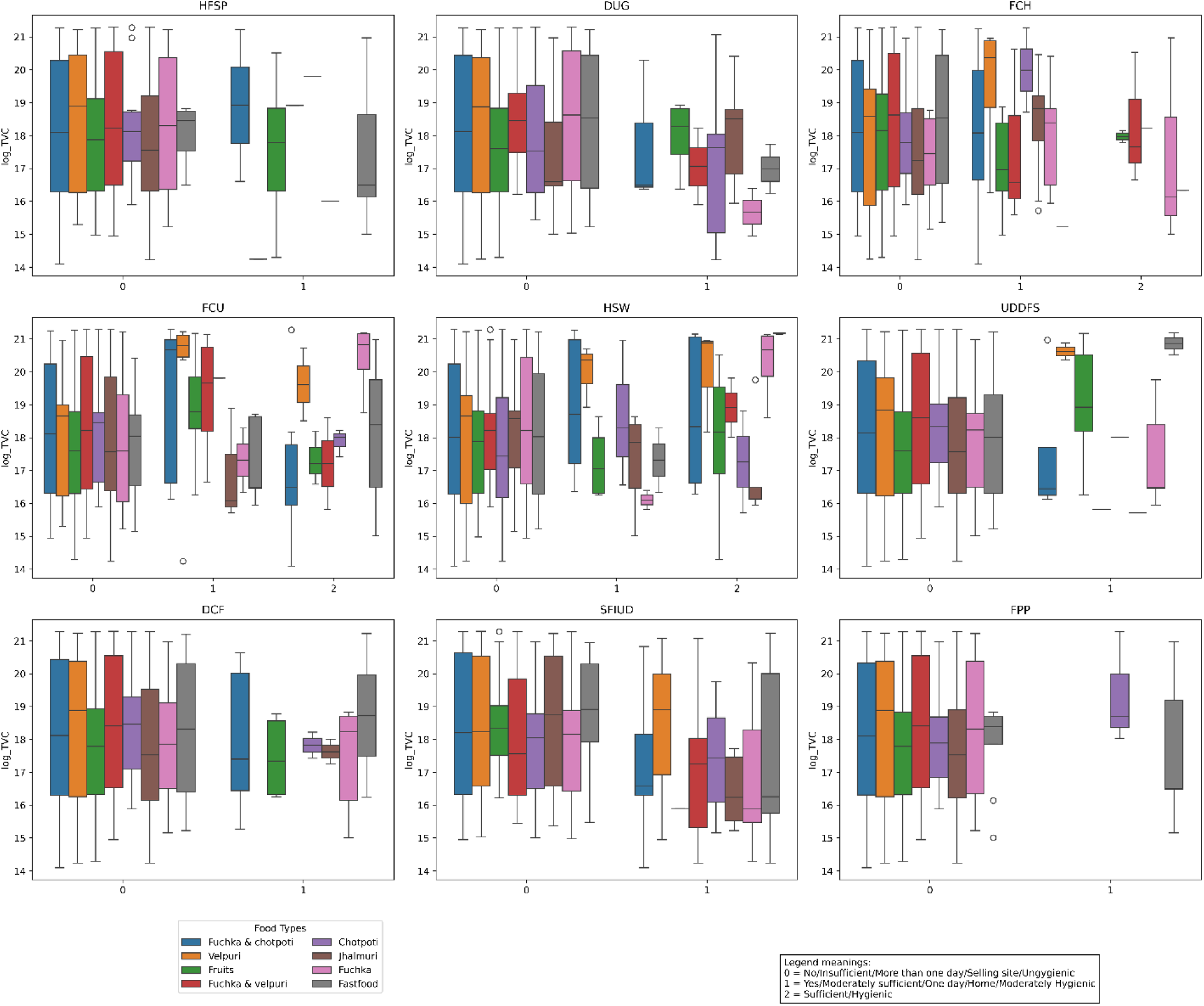
Variation of bacterial load in the food samples due to hygienic practices

**Table 2:**
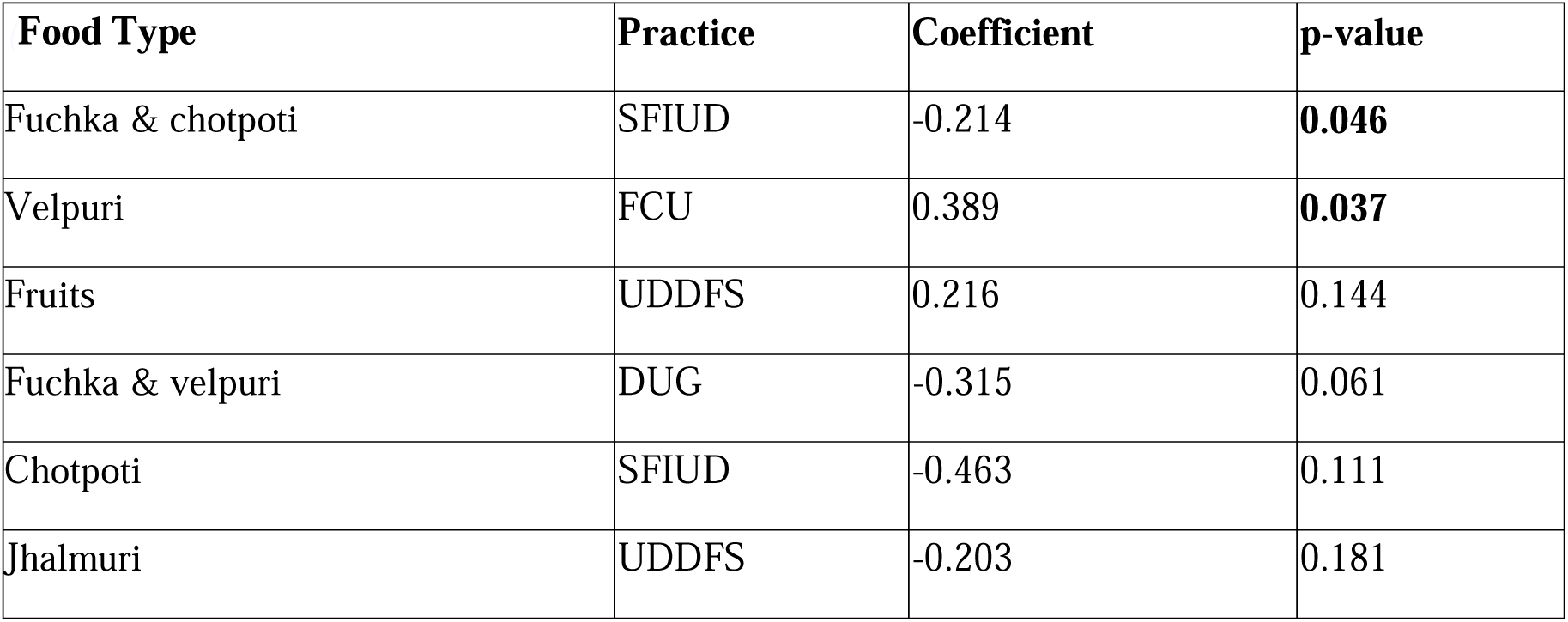
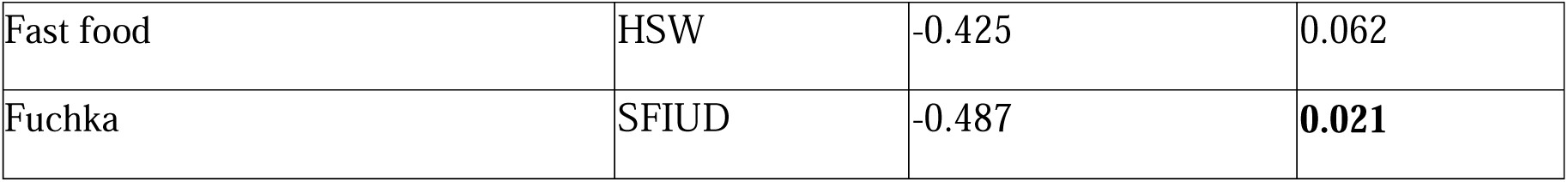
Spearman Correlation Results Between Hygiene Practices and Bacterial Contamination.

| Food Type | Practice | Coefficient | p-value |
| --- | --- | --- | --- |
| Fuchka & chotpoti | SFIUD | -0.214 | **0.046** |
| Velpuri | FCU | 0.389 | **0.037** |
| Fruits | UDDFS | 0.216 | 0.144 |
| Fuchka & velpuri | DUG | -0.315 | 0.061 |
| Chotpoti | SFIUD | -0.463 | 0.111 |
| Jhalmuri | UDDFS | -0.203 | 0.181 |

|  |  |  |  |
| --- | --- | --- | --- |
| Fast food | HSW | -0.425 | 0.062 |
| Fuchka | SFIUD | -0.487 | **0.021** |

### 3.7 Challenges for Hygienic Practices

The study revealed that street food vendors face key hygiene challenges, mainly high costs, knowledge gaps, and time constraints. Vendors without formal education were most affected by financial barriers (62.25%), while those with primary or secondary education reported more knowledge and time-related challenges, with 33.33% of primary-educated vendors citing time limitations. These findings, as shown in (Table 3) highlight the need for tailored interventions that address educational disparities and promote affordable solutions for improving food safety practices.

**Table 3:** Constraints for hygienic practices among the street vendors.

| **Educational Status** | **Higher Cost (%) [95% CI]** | **Knowledge Gaps (%) [95% CI]** | **Time Constraints (%) [95% CI]** | **Total Respondents** |
| --- | --- | --- | --- | --- |
| Uneducated | 62.25 [56.1–68.4] | 29.34 [23.6–35.1] | 8.41 [4.9–11.9] | 242 |
| Primary Education | 41.67 [27.9–55.4] | 33.33 [20.3–46.4] | 25.00 [12.8–37.3] | 48 |
| Secondary Education | 33.33 [4.4–62.3] | 50.00 [19.6–80.4] | 16.67 [0.0–38.9] | 10 |

## 4. Discussion

This study provides a comprehensive assessment of the hygienic practices of street food vendors in Mymensingh City, Bangladesh, highlighting key challenges and potential interventions to improve food safety. The findings show poor hygiene compliance shaped by education, financial limits, and weak regulation, echoing global street food safety studies and highlighting the need for targeted interventions to reduce foodborne risks (Kundu et al., 2021; Banna et al., 2022; Hossain and Habib, 2023). Education strongly influenced hygiene, with vendors holding secondary education demonstrating better food safety knowledge and practices than those without formal schooling, consistent with previous findings linking literacy to safer food handling (Kundu et al., 2021; Islam et al., 2024).

Financial constraints were identified as a significant barrier to proper hygiene compliance, particularly among uneducated vendors. Approximately 62.25% of uneducated vendors cited cost-related challenges, which is in line with previous studies that found economic limitations often deter vendors from adopting hygienic practices (Nizame et al., 2019). The expense of essential hygiene items can limit compliance among low-income vendors, emphasizing the need for subsidies or low-cost alternatives. Microbiological testing revealed high bacterial loads, with Velpuri showing the highest log-transformed total viable count, indicating elevated foodborne risk. Previous research has similarly documented high microbial contamination in street foods, attributing it to improper handling, lack of food covering, and inadequate sanitation (Hasan et al., 2021; Robeul Islam et al., 2024). The link between food safety knowledge and practices, such as glove use and food covering, underscores the importance of education-based interventions to reduce contamination. This is also proposed by the study reported by (Islam et al., 2024) which highlighted that greater food safety awareness is linked to better hygiene practices, like glove use and food covering, reducing contamination risk. A study similarly demonstrated that implementing stringent guidelines on ingredient freshness played a crucial role in minimizing microbial load in food samples (Gurnari, 2015). A study reported that antimicrobial-resistant strains of foodborne pathogens are increasing significantly worldwide due to climatic and socioeconomic factors, posing a serious concern for public health (Nahiduzzaman et al., 2026).

Although this study thoroughly assessed vendor hygiene and microbial contamination, its cross-sectional design limits causal conclusions. Observed links between knowledge and hygiene cannot confirm that education directly improves practices. Moreover, self-reported data may overestimate vendors’ compliance due to response bias (Adams et al., 1999). Future studies should employ longitudinal and randomized designs to evaluate hygiene interventions and consider consumer awareness alongside vendor practices to inform safer street food policies (Mukhola, 2014). Policies should mandate food safety training for low-education vendors, provide affordable sanitation, and enforce regulations with incentives. Collaboration between government, NGOs, and community programs can sustain improvements, reduce foodborne risks, and support the livelihoods of street vendors, a vital part of urban food systems in Bangladesh and similar settings.

## 5. Conclusion

This study reveals significant hygiene deficiencies among street food vendors in Mymensingh City, Bangladesh, including poor hygienic practices, inadequate food covering, and limited use of disinfectants. Substantial knowledge gaps and high bacterial contamination in some street foods highlight potential public health risks. Although education appears to improve hygienic practices, financial limitations and time constraints often hinder proper compliance. Therefore, targeted training programs, subsidized hygiene resources, stricter regulatory oversight, and community awareness initiatives are essential. Future longitudinal studies are recommended to evaluate the effectiveness of such interventions, while coordinated efforts among policymakers, health authorities, and vendors are crucial to ensure safer and more sustainable street food practices.

## 6. Ethics Statement

This study was conducted in accordance with established ethical standards for research involving human participants. Ethical approval was obtained from the Institutional Review Board of Bangladesh Agricultural University (Approval No: AWEEC/BAU/2024(2)/15(a)). All procedures performed in this study complied with the relevant institutional guidelines and regulations. Informed written consent was obtained from all participants prior to data collection, and confidentiality and anonymity were strictly maintained throughout the study.

## Data Availability

All data produced in the present study are available upon reasonable request to the authors

